# Food insecurity as a determinant of adolescent mental health in Francophone and Anglophone Africa: A multilevel analysis

**DOI:** 10.64898/2026.05.19.26353168

**Authors:** Cynthia Lum Fonta, Frank Elgar, David Gordon, Murray Leibbrandt, Zoi Toumpakari

**Affiliations:** South African Labour and Development Research Unit (SALDRU), University of Cape Town, Rondebosch, 7701, Cape Town, South Africa; School of Population and Global Health, McGill University, 2001 McGill College Avenue, Montreal, QC, H3A 1G1, Canada; School for Policy Studies, University of Bristol, 8 Priory Road, Bristol, BS8 1TZ, United Kingdom

**Keywords:** Food Insecurity, Mental Health, sub–Saharan Africa, Adolescence, Gallup Poll, Negative Experience Index

## Abstract

Food insecurity (also called, simply, FI) levels in sub-Saharan Africa are rising among its growing adolescent population, the world’s fastest-growing teenage population. This study examines food insecurity and its role as a social determinant of poor mental health among African adolescents. The study utilised the Gallup World Poll data between 2014 and 2019, including adolescents aged 15 -19 (n=25,368). Poor mental health was measured using five validated responses about negative experiences. We employed a two-level binary logistic regression model to determine the associations between food insecurity and poor mental health. At the individual level, the primary explanatory variable, food insecurity, was measured using the FAO (2015) Food Insecurity Experience Scale (FIES). The odds of poor mental health exhibited a dose-response relationship with food insecurity severity, with mild (OR=1.70; 95% CI (1.57-1.84), moderate (OR=2.35; 95% CI (2.17-2.54) and severe food insecurity (OR=3.19; 95% CI (2.96-3.54) being associated with poorer mental health. Other assessed covariates showed that residing in a Francophone state increased the odds of poor mental health experiences, whereas positive experiences reduced the chances of poor mental health, as did residing in rural areas. There was no difference in the relationship between mental health and food insecurity across the two colonial origins. Food insecurity remains an important determinant of adolescent mental health in Francophone and Anglophone Africa. Investing in cost-effective agricultural and nutrition-sensitive interventions that boost food production could improve adolescent mental well-being while reducing long-term social and economic burdens on families and health systems in sub-Saharan Africa.

## Introduction

Geographical and social patterns of mortality and morbidity are shaped not only by biological factors but also by a complex interplay of social, economic, political, and cultural influences (1). As a result, the social determinants of health framework provides a valuable context–process–outcome perspective for understanding these mechanisms, particularly in how material disadvantages contribute to poor health outcomes in global health research (2). Using this framework, this study examines how food insecurity affects mental health outcomes among adolescents in sub-Saharan Africa.

Adolescence (ages 10–19 years) represents a critical and vulnerable developmental stage characterised by rapid biological, emotional, and psychosocial transitions (3). Hormonal changes that enhance cognitive capacities and heightened social awareness render adolescents particularly susceptible to stressors that arise from household and societal adversity, including food insecurity (4). This vulnerability is especially concerning in sub-Saharan Africa, where adolescents constitute the fastest-growing section of the population (3), a trend driven mainly by regional inequalities in high fertility rates and unmet family planning services (5). Adolescent food insecurity can have significant consequences for both immediate and long-term physical and mental health, including anxiety, loneliness, suicidal ideation, suicidal plans and suicide attempts (6, 7). As food insecurity continues to increase across many African countries (8), the simultaneous growth of the adolescent population and chronic food deprivation raises significant concerns for mental well-being, human capital formation, and future productivity.

Despite this demographic and nutritional urgency, food insecurity and nutrition-related challenges among adolescents remain understudied across sub-Saharan Africa. Research and policy attention in low- and middle-income countries has historically prioritised early childhood and maternal nutrition, with adolescents receiving comparatively little focus in both research agendas and health interventions (9). This neglect is problematic, as adolescent-reported food insecurity offers unique insights into lived nutritional deprivation that may not be fully captured through parental information. Incorporating adolescents’ own perspectives is therefore essential for evidence-based policymaking that recognises adolescents as active social agents rather than passive dependants.

Studies worldwide consistently demonstrate that food insecurity is associated with heightened psychological distress, depression, anxiety, and suicidal behaviours among adolescents and young people (10–13). Research in Ghana (14), Ethiopia (12), Tanzania and Malawi (15) shows that inadequate access to food acts as a significant stressor, contributing to school dropout, early and unintended pregnancies, heightened anger, and increased exposure to gender-based violence. Notably, some studies reveal complex dynamics, such as higher psychological distress among adolescents in households receiving remittances, those living with HIV/AIDS and suggest that food insecurity interacts with broader social and economic vulnerabilities (16, 17).

The pathways through which food insecurity impacts adolescent mental health are complex. One key mechanism is biological. Food deprivation reduces intake of essential micronutrients such as iron, zinc, folate, and vitamin B12, which are critical for neurotransmitter synthesis, brain oxygenation, and emotional regulation (18, 19). Anaemia, a consequence of micronutrient deficiency, reduces physical activity and hampers cognitive development, which can negatively affect academic performance and future prospects (20). Beyond biological effects, food insecurity also has psychosocial impacts among adolescents (21). Experiences of hunger may lead to feelings of shame and social exclusion, particularly among individuals who rely on food aid or live in impoverished environments (22, 23). Research in Ethiopia and Brazil indicates that food insecurity is a basic needs deprivation that may have a direct effect on mental health by causing stress (e.g. worry about where the next meal is coming from, etc.) (24). Evidence also indicates that absolute food insecurity is more strongly linked to poor mental health than relative deprivation, particularly in settings with high prevalence where social comparison and related stigma may be less pronounced (25). Seasonal variations further influence this relationship: in rural Tanzania, mental health outcomes worsened during periods of increased food insecurity in the rainy season and improved after harvest in the dry season (26).

This study also explores other socio-economic and political determinants of poor mental health aside from food insecurity. For example, Bjornlund et al. argue that colonial agricultural systems in some parts of sub-Saharan Africa prioritised export-oriented production for Global North markets, diverting land, labour, and resources away from domestic food production and weakening traditional food security systems (27). These colonial legacies, sustained through post-independence policies and structural economic constraints, have limited investment in local food systems and contributed to persistent food insecurity across the region. Other studies have linked the rise of poor mental health to urbanisation (28), income instability (29) and the absence of social support systems, such as remittance (30). These factors will be controlled for in this analysis.

Mental health remains under-prioritised in sub-Saharan Africa. Limited data availability, cultural stigma surrounding mental illness, and underinvestment in mental health infrastructure have constrained policy recognition and service provision across the region. As Glover-Thomas and Chima note, mental health typically appears last on national development agendas, with patients in low-resource settings often relying on out-of-pocket payments to access care (31). Such neglect is particularly alarming given that at least 20% of the world’s children and adolescents experience some form of mental health disorder before the age of 40 years (32). This cross-national evidence shows that food insecurity is an important and preventable risk factor for poor mental health outcomes in Africa. Interventions that reduce food insecurity are among the most cost-effective ways to lift people out of hunger (33) and, in the long term, prevent the economic burden of mental health illnesses to the state and family (34).

The mental health-food insecurity relationship is well-documented in rich countries (35, 36) and most parts of Anglophone Africa (37, 38) but little is known in Francophone states. This research gap is concerning, given that several conflict-affected Francophone nations (the Central African Republic, Chad, the Democratic Republic of the Congo, and Madagascar) are among the countries with the highest levels of hunger worldwide (39). Yet, the mental health consequences of ongoing food deprivation in these regions remain underexplored. This research investigates the relationship between food insecurity and adolescent mental health across both Francophone and Anglophone African countries, while controlling for other poor mental health covariates.

## Materials and Methods

This analysis used secondary data from the Gallup World Poll (GWP), which surveyed adolescents, young adults, and older adults (>15 years) in over 140 countries since 2005. The anonymised dataset was accessed for research purposes in June 2020, and the authors did not have access to any personally identifiable information during or after data collection. The poll included questions across various domains and indicators, such as mental health experiences and food insecurity. This research focuses on low, middle, and upper-middle-income countries in sub-Saharan Africa, which bear a high burden of food insecurity (8). Second, the country selection was limited to Francophone and Anglophone states previously colonised by France and the United Kingdom to examine the relationship between food insecurity and mental health in these two ex-colonial contexts. Third, the unit of analysis comprised only adolescents aged 15–19 years.

Table 1 presents sample statistics of adolescents representing 34 African states, 15 Francophone states (52%), and 19 Anglophone states (48%), interviewed between 2014 to 2019 (i.e. before the Covid pandemic). The sample includes a total of 25,363 adolescents in sub-Saharan Africa, with the smallest sample in Sudan and the highest in Niger.

**Table 1:**
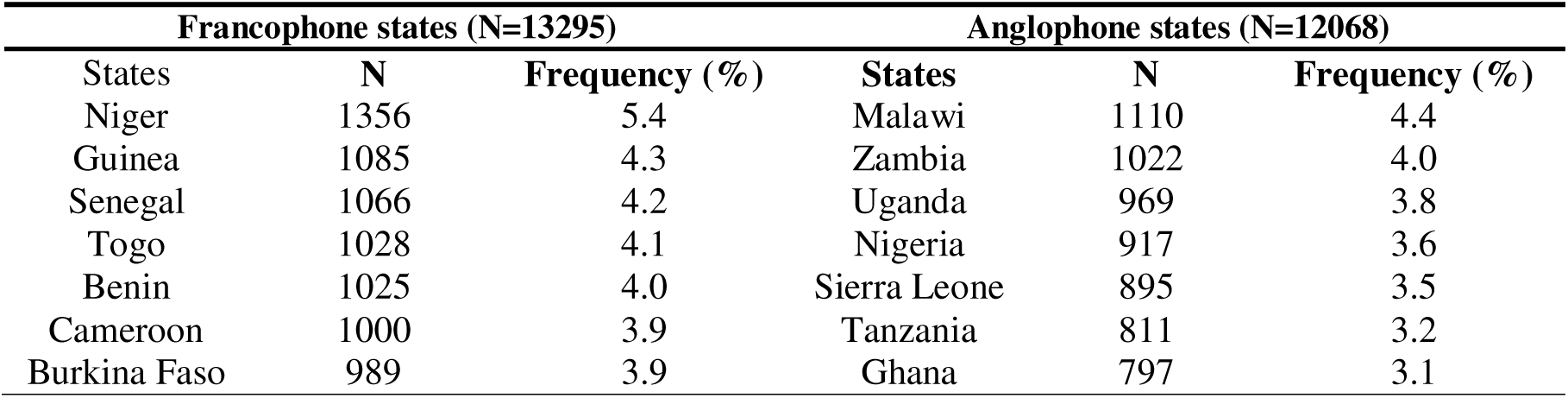
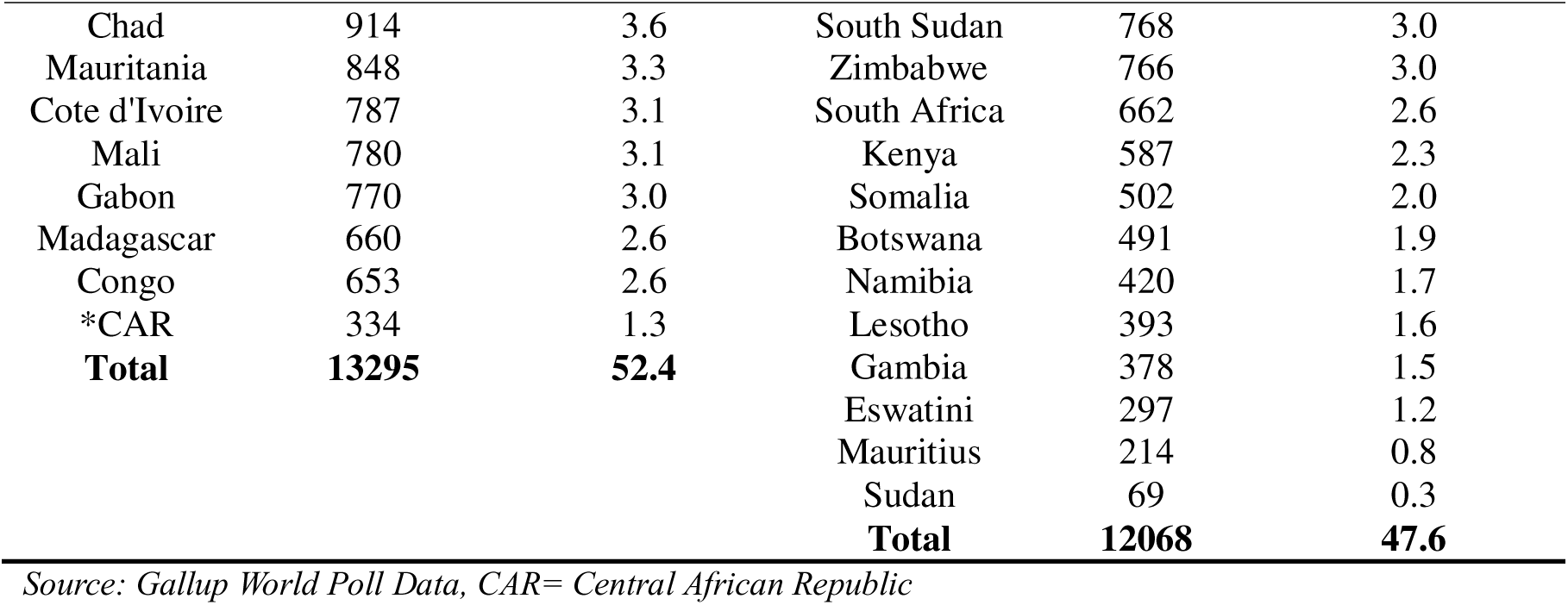
Unweighted sample distribution of adolescents in Sub-Saharan Africa (2014–2019)

### Key variable measures

The outcome variable, the negative well-being index, serves as a proxy for poor mental health. Adolescents reported the following adverse experiences the previous day: feelings of physical pain, worries, sadness, stress, and anger. Responses were coded as one for affirmative answers and zero otherwise. The internal consistency of these five items within this age group was 0.7 (Cronbach’s alpha). Affirmative responses to all five negative experiences were summed to form a scale from 0 to 5, where 0 indicates none of the experiences, and 5 indicates all are present. This five-point scale constitutes the negative experience index (NEI), which was dichotomised into ‘0’ for no negative experience and ‘1’ for at least one negative experience. The negative experience index score was used as a measure of poor mental health. Previous studies have used the NEI as a proxy for poor mental health to examine its association with food insecurity (25, 40).

This study’s primary independent variable, food insecurity, was measured using the Food Insecurity Experience Scale (FIES). The scale was developed by the Food and Agriculture Organisation (FAO) Voices of the Hungry Project (VOH) (41). The FIES provides the most suitable and valid indicators of food insecurity for monitoring Sustainable Development Goal 2. Food insecurity is assessed on a scale of 8 items, ranging from mild to severe. Individuals with the highest raw score of eight had the highest probability of being food insecure, and those with the lowest score had the lowest likelihood of being food insecure.

Table 2 presents weighted and unweighted proportions of yes responses to the FIES questions. Most respondents reported less severe forms of food insecurity (e.g., worrying about food, having no healthy meals, eating fewer foods or skipping meals), and fewer reported the most severe form (a whole day without food).

**Table 2:** The proportion of respondents’ food insecurity experience items.

| Food insecurity experience items (code names) | % Unweighted<br>(N=20,007) | % Weighted<br>(N= 24,247) |
| --- | --- | --- |
| Experience of worry (Worried) | 59 | 60 |
| Experience of no health foods (Healthy) | 58 | 59 |
| Experience of fewer foods (Fewer foods) | 62 | 63 |
| Experience of Skipping meals (Skipped) | 54 | 55 |
| Experience of eating less (Ate less) | 61 | 62 |
| Experience of running out of food (Run out) | 50 | 52 |
| Experience of going hungry (Hungry) | 51 | 52 |
| Experience a day without eating (Whole day) | 35 | 36 |

### Observations for only the yes responses depicting food insecurity

Following the FAO methodology, equating plots were used to identify comparable item estimates across states and indicated that the ‘Ate less’ question responses deviated very slightly from all others. Dropping the ‘Ate less’ question responses improved the correlation between the items from 92% to 95%. Given the high correlation estimates for both the seven items (95%) and all eight items (92%) of the instrument, no items were dropped when calculating food insecurity prevalence. Adolescents with FIES scores of 1-3 were classified as suffering from mild food insecurity, 4-6 as moderate, and 7-8 as severe.

All analyses were conducted using STATA version 18 and SPSS Statistics version 26 for descriptive statistics. Descriptive statistics were provided for the study population, including country-level characteristics, population sizes, and socioeconomic and demographic features. The means and standard deviations of essential variables were presented by colonial origin. Furthermore, Spearman’s correlation (Rho) matrix was used to identify significant associations for regression analysis. Lastly, we estimated a two-level mixed-effects model to examine how individual- and country-level factors explain mental health variability, as specified:

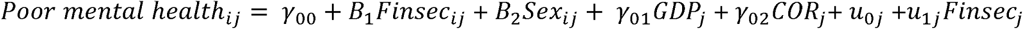

Where

|  |  |
| --- | --- |
| $\gamma_{00}$ | Grand mean intercept (fixed effect) |
| $B_1$ and $B_2$ | Individual-level covariate coefficients |
| $\gamma_{01}$ and $\gamma_{02}$ | country-level covariates coefficients |
| $u_{0j}$ | country-specific random intercept |

### Random part

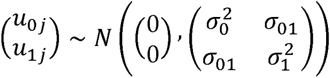

Interpretation:

- 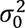: between-country variance in baseline poor mental health (random intercept)
- 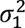: between-country variance in the effect of food insecurity (random slope)
- *σ*_01_: covariance between baseline risk and FI effect (do higher-baseline countries also have stronger/weaker FI effects?)

Individual-level covariates (food insecurity, positive experience index (PEI), age, sex, education, remittance and income per capita) were specified at level 1. In contrast, country-level structural factors (GDP per capita and colonial origin) were specified at level 2 (See Table 3 below). The model also incorporates random-slope variation, allowing both baseline mental health risk and the effect of food insecurity to vary across countries. This controls for unobserved country-specific heterogeneity.

**Table 3:** Variable table.

| Variables | Description | Measurement |
| --- | --- | --- |
| **Dependent variable** | Negative Experience Index (NEI) Scale, | Scaled as an ordinal variable from 0 – 5. At least one negative experience depicted poor mental health experience |
| **Independent variable** |  |  |
| **Food insecurity** | Respondents were asked if they experienced any of the eight items in the FIES | 1-3 items= mild FI<br>4-6 items = moderate FI<br>7-8 items= severe FI |
| **Individual-level covariates** |  |  |
| **Income groups** | Income= All incoming wages after exclusion of taxes, remittances | The income variable was classified into five quintiles ranging from lowest to highest income (Q1-Q5) |
| **Positive experience index** | Positive Experience Index Scale (PEI) | Scaled as an ordinal variable from 0 – 5, with five depicting the highest positive mental health state |
| **Education level** | This was classified into no education, 1-8 years of primary, secondary, and post-secondary education. | No basic education category was coded “1”; basic, secondary, and tertiary education categories were grouped and coded “0”. |
| **Gender** | Male versus female | Males coded “0” and females “1” |
| **Age** | Adolescents 15-19 years | Continuous variable |
| **Place of residence** | Rural vs urban | Rural residents coded “0” and urban residents “1” |
| **Country-level covariates** |  |  |
| **Country GDP** | Gross Domestic Product per Capita 2021 World Bank grouping | Continuous variable |
| **Colonial origin** | Francophone vs Anglophone states | Francophone states=1, Anglophone states=0 |

Source: Gallup (2017)

The null (empty) model provides two key pieces of information. First, it estimates the overall baseline level of poor mental health across countries, expressed as the average log-odds for individuals in an average country. Second, it quantifies the between-country variance in poor mental health, which indicates the extent to which outcomes cluster at the country level. This variance informs whether a multilevel specification is warranted. In this analysis, the intraclass correlation coefficient (ICC) was 0.06, indicating that approximately 6% of the total variance in poor mental health was attributable to between-country differences. This exceeds the commonly used threshold of 5%, demonstrating meaningful non-independence of observations within countries and justifying the use of a multilevel modelling approach. Likelihood-ratio tests comparing the multilevel model with a single-level logistic regression further supported this choice (χ² = 854.8, p < 0.001), indicating that the multilevel model provided a significantly better fit to the data.

### Postestimation effects

Marginal effect provide quantifiable estimates of the effects of food insecurity on mental health for Francophone and Anglophone states. This entailed calculating marginal effects from post-logistic regressions. The marginal effects estimate the differences in food insecurity means and their effects on poor mental health across the two colonial origins.

## Results

Table 4 shows weighted descriptive socio-demographic features of adolescents. The average age of adolescents was 17 years (SE 1.7), with 51% boys and 49% girls. Of these children, 69% lived in rural areas, and over 84% were attending primary, secondary, or tertiary institutions. Out of the total adolescents, 31% received remittances from relatives abroad, and more (22%) belonged to the low-per-capita-income group, compared with 18% in the richest income quintile.

**Table 4.** Weighted sociodemographic characteristics.

| **Variables** | **Weighted**<br>**N=30,448** | **Percentage (%)** |
| --- | --- | --- |
| **Sex** |  |  |
| Boys | 15429 | 51 |
| Girls | 15019 | 49 |
| **Age (years)** |  |  |
| 15 | 6462 | 21 |
| 16 | 5740 | 19 |
| 17 | 5808 | 19 |
| 18 | 6906 | 23 |
| 19 | 5533 | 18 |
| **Locality** |  |  |
| Urban | 9471 | 31 |
| Rural | 20807 | 69 |
| **Education** |  |  |
| None | 4915 | 16 |
| Primary | 12477 | 41 |
| Secondary/tertiary | 13056 | 43 |
| **Income quintile** |  |  |
| Poorest 20% | 6483 | 22 |
| Second 20% | 6221 | 21 |
| Middle 20% | 5925 | 20 |
| Fourth 20% | 5742 | 19 |
| Richest 20% | 5372 | 18 |
| **Remittance** |  |  |
| No | 17604 | 69 |
| Yes | 7977 | 31 |
| **Colonial origin** |  |  |
| Francophone states | 15481 | 51 |
| Anglophone states | 14967 | 49 |
| **Variables** | **Mean (SE)** | **95% CI** |
| Mean age (years) | 17(1.4) | (16.9-17) |
| Mean GDP per capita (PPP dollars) | 1846 (11) | (1824-1867) |

Table 5 (below) presents summary measures of adolescent mental health experiences across sub-Saharan Africa. Worrying the previous day was the most frequently reported negative experience (M=0.35, SD=0.48), and anger (M=0.22, SD=0.41) was the least reported. Overall, most adolescents reported positive experiences the previous day, particularly feeling respected (M=0.80, SD=0.40) and learning something new (M=0.64, SD=0.48).

**Table 5.** Weighted summary statistics of mental health experiences yesterday.

| Variable | Mean | Standard<br>Deviation | 95% CI |
| --- | --- | --- | --- |
| **Negative experiences** |  |  |  |
| Worry | 0.35 | 0.48 | (0.34, 0.35) |
| Pain | 0.31 | 0.46 | (0.30, 0.31) |
| Stress | 0.28 | 0.45 | (0.27, 0.29) |
| Sadness | 0.24 | 0.43 | (0.24, 0.25) |
| Anger | 0.22 | 0.41 | (0.21, 0.22) |
| **Positive experiences** |  |  |  |
| Respected | 0.80 | 0.40 | (0.79, 0.81) |
| Smiled | 0.78 | 0.41 | (0.77, 0.79) |
| Well-rested | 0.72 | 0.45 | (0.71, 0.72) |
| Enjoyment | 0.65 | 0.48 | (0.64, 0.65) |
| Learned something new | 0.64 | 0.48 | (0.63, 0.65) |

243 *Sample = 30,448 (weighted)*

Table 6 (below) presents the overall prevalence of food insecurity (independent variable) and poor mental health (outcome variable) by Francophone and Anglophone countries. There were no statistically significant differences in food insecurity and poor mental health between the two ex-colonial origins. However, adolescents in Anglophone states experienced more severe food insecurity (42%) than Francophone adolescents (32%), while mild to moderate food insecurity was more prevalent in Francophone states. Overall, 37% of adolescents in sub-Saharan Africa were living with severe food insecurity, 24% in the moderate category and 19% with mild food insecurity.

**Table 6.** Univariate analysis of Negative Experience Index and risk factors by state type.

| Variable | Francophone<br>state (%)<br>N=15,481 | Anglophone<br>state (%)<br>N=14,967 | Risk ratios<br>(95% CI) |
| --- | --- | --- | --- |
| **Food insecurity** |  |  |  |
| None | 20 | 20 | 0 |
| Mild food insecurity | 22 | 16 | 1.2 (0.9, 1.6) |
| Moderate food insecurity | 25 | 22 | 1.1 (0.8, 1.5) |
| Severe food insecurity | 32 | 42 | 0.8 (0.6, 1.1) |
| **Negative experience scale** |  |  |  |
| 1 | 24 | 22 | 1.1 (0.8, 1.5) |
| 2 | 18 | 15 | 0.8 (0.5, 1.2) |
| 3 | 12 | 10 | 1.1 (0.7, 1.8) |
| 4 | 8 | 7 | 1.1(0.6, 1.7) |
| 5 | 4 | 5 | 0.9 (0.4, 1.9) |
| **NEI** |  |  |  |
| None | 34 | 42 | 0.8 (0.6, 1.1) |
| At least one negative experience | 66 | 58 | 1.2 (0.9, 1.7) |
| **Positive experience scale** |  |  |  |
| 0 | 3 | 3 | 0 |
| 1 | 7 | 6 | 1.1 (0.6, 1.8) |
| 2 | 12 | 10 | 1.1 (0.7, 1.7) |
| 3 | 20 | 17 | 1.1 (0.8, 1.5) |
| 4 | 28 | 28 | 0 |
| **PEI** |  |  |  |
| 0 to 4 positive experiences | 72 | 64 | 1.2 (0.9, 1.7) |
| All five positive experiences | 28 | 36 | 0.8 (0.6, 1.1) |

**NEI: Negative Experience Index, PEI: Positive Experience Index**

Most adolescents reported fewer negative experiences and more positive experiences. Forming a binary variable from the experience scale, 62% of adolescents reported at least one negative experience. Positive experiences were treated as an independent variable, with 28% of Francophone children reporting all five experiences compared with 36% of Anglophone children.

Table 7 presents a sequence of mixed-effects two-level regression models that analyse the association between food insecurity and poor mental health among adolescents, while accounting for both individual- and country-level factors. The null model estimated a fixed-effects intercept of 0.44 (95% CI: 0.23–0.62), representing the average log-odds of poor mental health among adolescents across countries. When converted to the probability scale, this corresponds to an overall prevalence of poor mental health of about 61%, aligning with descriptive estimates and confirming a significant baseline mental health burden across the study population.

**Table 7.**
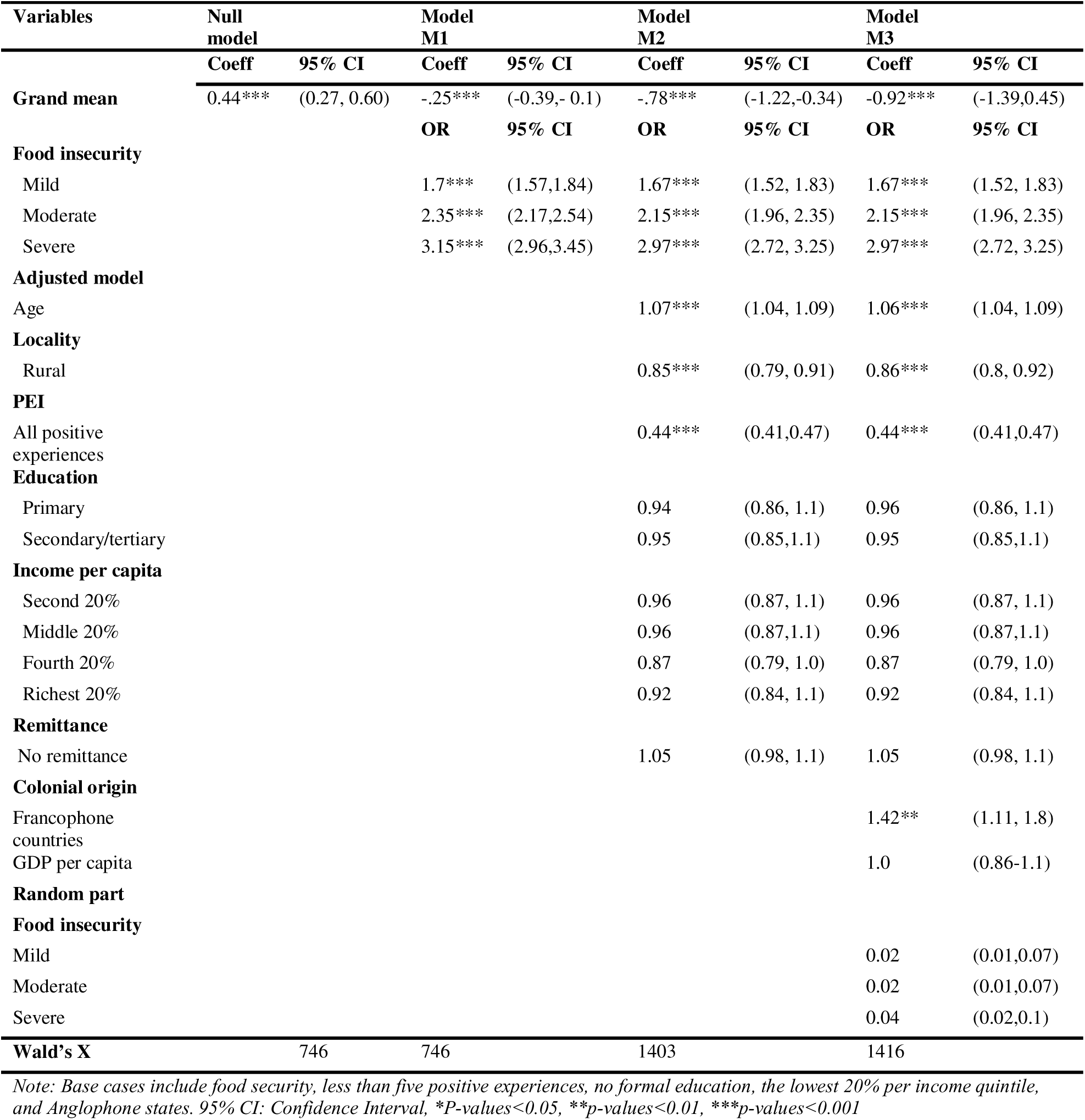
Two-level regression models of food insecurity and poor mental health.

Model M1 introduces food insecurity at the individual level and reveals a clear dose–response relationship. Relative to food-secure adolescents, the odds of poor mental health increased progressively with the severity of food insecurity. That is, 1.7 times (OR=1.70; 95% CI (1.57-1.84) higher among mildly food-insecure adolescents, 2.4 times higher among those moderately food-insecure (OR=2.35; 95% CI (2.17-2.55), and more than threefold higher among severely food-insecure adolescents (OR=3.19; 95% CI (2.95-3.45). This graded pattern indicates a strong and monotonic association between worsening food access and deteriorating mental health.

Model M2 further adjusts for key individual-level covariates, including age, place of residence, positive mental health experiences, education, household income, and remittances. Older age was associated with higher odds of poor mental health, with each additional year increasing the risk by approximately 7% (OR=1.07, p < 0.001). Adolescents residing in rural areas were 15% less likely to report poor mental health compared with their urban counterparts (OR = 0.85, p < 0.001). Notably, adolescents reporting all five positive mental health experiences had substantially lower odds of poor mental health (OR = 0.44, p < 0.001), highlighting the protective role of positive experiences in this age group. Importantly, the magnitude of the food insecurity coefficients changed minimally after adjustment, suggesting that the association between food insecurity and poor mental health is largely independent of these individual-level socioeconomic and demographic factors.

Model M3 incorporates country-level covariates to capture broader structural influences. Adolescents residing in Francophone countries exhibited significantly higher odds of poor mental health compared with those in Anglophone countries (OR = 1.4, p < 0.05), highlighting the relevance of historical and institutional contexts. In contrast, GDP per capita was not significantly associated with poor mental health. Suggesting that higher national economic wealth alone may not necessarily translate into better adolescent mental health outcomes. The food insecurity coefficients remained stable, underlining their strong association with poor mental health, even after adjusting for individual and country-level factors.

Across models, the progressive decline in the intercept reflects decreasing baseline risk among reference food-secure adolescents as explanatory covariates are introduced. The random part of the model showed no significant heterogeneity in the association between food insecurity and poor mental health across countries, suggesting that although baseline mental health risk varies by country, the strength of the food insecurity–mental health relationship is broadly consistent across national contexts. That is, food insecurity increases poor mental health everywhere, but it does so by roughly the same amount in all countries.

Figure 1 illustrates the relationship between food insecurity and poor mental health by outlining the predictive probabilities of poor mental health outcomes and food insecurity among adolescents. The predictive probabilities quantify the odds of experiencing poor mental health given food insecurity. Poor mental health experiences increased with the increasing severity of food insecurity. The probabilities of poor mental health (47%) were lowest amongst food-secure adolescents and consistently increased in the mild (58%), moderate (63%) and severe (70%) categories of food-insecure adolescents.

Figure 2 presents differences between food-insecure and food-secure adolescents, with poor mental health experiences, which were also compared between Francophone and Anglophone states. The marginal effects show a strong dose–response relationship between food insecurity and poor mental health in both Francophone and Anglophone countries, with risk increasing sharply as food insecurity worsens. However, the overlapping confidence intervals suggest no statistically meaningful difference between the two groups of countries.

## Discussion

This analysis has shown that greater severity of food insecurity correlates with an increased risk of poor mental health among adolescents in sub-Saharan African countries. This association was not affected by whether adolescents resided in an Anglophone or Francophone country, emphasising the importance of ensuring food security across all sub-Saharan African countries. This finding contributes to the existing literature on the socioeconomic determinants of poor mental health in sub-Saharan Africa. To effectively prevent mental health issues, it is essential to ensure that no child goes hungry, especially in Francophone countries, where even mild food insecurity can significantly elevate the risk of poor mental health.

Our research findings are consistent with Elgar and colleagues (2021) study of 160 countries, which used similar measures of poor mental health. They found strong associations between poor mental health and absolute food insecurity in areas where absolute food insecurity was high (25). However, with respect to relative food insecurity (i.e., food insecurity associated with income), the authors observed a stronger association between relative food insecurity and poor mental health in areas with lower food insecurity prevalence. Food insecurity in high-income areas may trigger mental health experiences among food-insecure individuals, based on the relative deprivation theory of poverty (42).

Townsend’s theory of poverty may offer a plausible explanation as to why rural adolescents were less likely to report poor mental health experiences than their urban counterparts in this analysis. In a deprived community, where everyone lacks material needs, there might be fewer poor mental health experiences from feeling socially excluded. The sense of shame and humiliation may be reduced when many people undergo similar experiences. Our findings, however, contradict studies that associate rural areas with poor mental health experiences (43, 44).

Adolescents in Francophone countries exhibited a higher probability of poor mental health compared with their Anglophone counterparts. Identifying the specific drivers of cross-national differences in adolescent mental health is challenging, particularly because state-level factors in this analysis accounted for only a small proportion of the overall variation relative to individual-level characteristics. This challenge is compounded by the limited availability of reliable mental health prevalence data in many Francophone African countries, which constrains the identification of plausible explanatory pathways (45).

Nevertheless, some evidence suggests that the widespread presence of low-quality schools attended predominantly by socioeconomically disadvantaged students in several Francophone countries may adversely affect academic performance (46). Such educational environments may also affect adolescents’ mental well-being, particularly in high-pressure contexts such as national academic competitions. In addition, the high prevalence of conflict and insecurity in many Francophone countries, including Niger, Mali, Cameroon, Burkina Faso, and the Central African Republic, may further exacerbate poor mental health outcomes during adolescence. These dynamics warrant further empirical investigation, particularly regarding the role of violence and conflict exposure in shaping adolescent mental health in this region.

We found no significant variation in poor mental health across income groups. This likely reflects limitations in income measurement, especially among adolescents who do not earn an independent income and may report household economic status imprecisely.

### Strengths and limitations

This study has several limitations. First, the cross-sectional and cross-national nature of the data preclude causal inference about the relationship between food insecurity and adolescent mental health. Second, mental health was measured using self-reported emotional experiences rather than clinically diagnosed conditions such as anxiety or depression, which are among the most prevalent mental health disorders during adolescence. Third, responses may be subject to cultural interpretation and reporting biases across diverse country contexts. Nonetheless, the emotional experience indices used in this study have demonstrated high reliability and construct validity in the Gallup World Poll, supporting their use as reasonable proxies for population-level mental health outcomes. However, both food insecurity and mental health are complex phenomena (47), and there are many factors that are associated with them that we have not been able to control for in this analyses (e.g. bullying at school, etc.). This analysis, may therefore, suffer from some omitted variable bias.

Despite these limitations, the study has notable strengths. The large sample size and inclusion of more than half of sub-Saharan African countries enhance statistical power, improve external validity, and allow for robust cross-country comparisons, thereby strengthening confidence in the observed associations.

### Implications for research and policy

Further research on the mental health implications of food insecurity in Francophone Africa is warranted, given the relative scarcity of evidence from this context. Expanding research capacity and dissemination in the region through research funding and advocacy would strengthen the global evidence base and inform more context-responsive policy solutions. The evidence generated in this study supports the need for a more integrated approach to food security, one that explicitly incorporates mental health considerations within nutritional, agricultural, and social protection interventions across sub-Saharan Africa. For example, Ethiopia’s Productive Safety Net Programme integrates food assistance, livelihood support, and nutrition interventions, demonstrating that strengthening household food security can simultaneously reduce economic stressors associated with poor mental health outcomes. Nutrition-sensitive agriculture programmes implemented across sub-Saharan Africa by the Food and Agriculture Organisation and International Food Policy Research Institute link, diversified crop production, women’s empowerment, and dietary diversity to improve child and adolescent well-being.

Again, the study highlights the need to include valid mental health and food insecurity question modules in the DHS, MICS and other nationally representative social surveys. This addition would enable improved mental health needs monitoring of children and adolescents in sub-Saharan Africa. Finally, given the observed increase in poor mental health with greater severity of food insecurity, global and national actors should prioritise further research and interventions in conflict-affected regions and areas experiencing intensifying climate shocks, where disruptions to food systems are most acute. Targeting these high-risk settings is essential for mitigating both food insecurity and its downstream mental health consequences across the population in general and not just adolescents.

### Conclusion

Adolescents represent a critical yet frequently overlooked population in public health planning across sub-Saharan Africa. This study contributes evidence to address this gap by highlighting the mental health vulnerabilities of adolescents and nutritional factors shaping their well-being. Adolescent health is foundational to the future productivity and social development of African countries, yet many governments continue to under-invest in adolescent-focused services and in mental health systems more broadly. Health systems across the region remain under-resourced in the provision of counselling, diagnosis, and treatment, limiting timely prevention and care.

Against this backdrop, our findings underline food insecurity as a key and preventable determinant of poor mental health among adolescents. The strong, graded association observed suggests that addressing food insecurity is likely to be a public health strategy for reducing mental health prevalence in this age group. Preventive approaches that strengthen food security may generate significant mental health benefits, especially in resource-constrained settings where treatment-based services are limited. Investments in best practices that improve agricultural productivity and scale up nutrition-sensitive programmes can enhance dietary adequacy, strengthen household resilience, and mitigate upstream risk factors associated with poor mental health outcomes.

## Data Availability

The data is available upon institutional approval.

## Declaration section

## Acknowledgement

This work is a part of my doctoral thesis. I would like to acknowledge funding from the South-West Doctoral Training Partnership at the University of Bristol, which supported me in completing my programme. I also wish to express my gratitude to Helen Gordon for her support and valuable guidance in editing the manuscript.

## Ethical clearance

The School for Policy Studies Research Ethics Committee, University of Bristol, reviewed the ethical implications of this research and provided ethical approval.

## Notes

### Competing Interest Statement

The authors have declared no competing interest.

### Funding Statement

This work is a part of my doctoral thesis. I want to acknowledge funding from the South-West Doctoral Training Partnership at the University of Bristol, which supported me in completing my programme.

### Author Declarations

The School for Policy Studies Research Ethics Committee, University of Bristol, reviewed the ethical implications of this research and provided ethical approval for this work.

